# Applications of Natural Language Processing at Emergency Department Triage: A Systematic Review

**DOI:** 10.1101/2022.12.20.22283735

**Authors:** Jonathon Stewart, Juan Lu, Adrian Goudie, Glenn Arendts, Shiv A Meka, Sam Freeman, Katie Walker, Peter Sprivulis, Frank Sanfilippo, Mohammed Bennamoun, Girish Dwivedi

**Affiliations:** School of Medicine. The University of Western Australia, Crawley, Western Australia, Australia; Harry Perkins Institute of Medical Research, Murdoch, Western Australia, Australia; Department of Emergency Medicine, Fiona Stanley Hospital, Murdoch, Western Australia, Australia; Department of Computer Science and Software Engineering, The University of Western Australia, Crawley, Western Australia, Australia; HIVE & Data and Digital Innovation, Royal Perth Hospital, Perth, Western Australia, Australia; Department of Emergency Medicine, St Vincent’s Hospital Melbourne, Melbourne, Victoria, Australia; SensiLab, Monash University, Melbourne, Victoria, Australia; School of Clinical Sciences at Monash Health, Monash University, Melbourne, Victoria, Australia; Western Australia Department of Health, East Perth, Western Australia, Australia; School of Population and Global Health, University of Western Australia, Crawley, Western Australia, Australia; Department of Cardiology, Fiona Stanley Hospital, Murdoch, Western Australia, Australia

## Abstract

**INTRODUCTION:** Millions of patients attend emergency departments (EDs) around the world every year. Patients are triaged on arrival by a trained nurse who collects structured data and an unstructured free-text history of presenting complaint. Natural language processing (NLP) uses various computational methods to analyse and understand human language, and has been applied to data acquired at ED triage to predict various outcomes. The objective of this systematic review is to evaluate how NLP has been applied to ED triage, assess if NLP based models outperform humans or current risk stratification techniques, and assess if incorporating free-text improve predictive performance of models when compared to predictive models that use only structured data.

**METHODS:** All English language peer-reviewed research that applied an NLP technique to free-text obtained at ED triage was eligible for inclusion. We excluded studies focusing solely on disease surveillance, and studies that used information obtained after triage. We searched the electronic databases MEDLINE, Embase, Cochrane Database of Systematic Reviews, Web of Science, and Scopus for medical subject headings and text keywords related to NLP and triage. Databases were last searched on 01/01/2022. Risk of bias in studies was assessed using the Prediction model Risk of Bias Assessment Tool (PROBAST). Due to the high level of heterogeneity between studies, a metanalysis was not conducted. Instead, a narrative synthesis is provided.

**RESULTS:** In total, 3584 studies were screened, and 19 studies were included. The population size varied greatly between studies ranging from 1.8 million patients to 762 simulated encounters. The most common primary outcomes assessed were prediction of triage score, prediction of admission, and prediction of critical illness. NLP models achieved high accuracy in predicting need for admission, critical illness, and mapping free-text chief complaints to structured fields. Overall, NLP models predicted admission with greater accuracy than emergency physicians, outperformed abnormal vital sign trigger and triage score at predicting critical illness, and were more accurate than nurses at assigning triage scores in two out of three papers. Incorporating both structured data and free-text data improved results when compared to models that used only structured data. The majority of studies were (79%) were assessed to have a high risk of bias, and only one study reported the deployment of an NLP model into clinical practice.

**CONCLUSION:** Unstructured free-text triage notes contain valuable information that can be used by NLP models to predict clinically relevant outcomes. The use of NLP at ED triage appears feasible and could allow for early and accurate prediction of multiple important patient-oriented outcomes. However, there are few examples of implementation of into clinical practice, most research in retrospective, and the potential benefits of NLP at triage are yet to be realised.

## INTRODUCTION

Millions of patients attend emergency departments (EDs) around the world every year.^1^ Queues for care are common, so patients are often triaged on arrival to the ED by a trained nurse. Triage is central to the practice of emergency medicine.^2^ In the face of excess demand, triage allows EDs to allocate their finite resources in an equitable, efficient, and standardised way.^3,4^ Triage systems in current use include the Emergency Severity Index (ESI), Australasian Triage Scale (ATS), Manchester Triage Scale (MTS), and the Korean Triage and Acuity Scale (KTAS).^3,5^ Triage systems aim to aid emergency care providers in making a structured decision regarding the urgency of care that a patient requires, and in doing so, identify and prioritise those patients with time-sensitive care needs.^3,4^ However, urgency of care does not necessarily reflect severity of illness (as judged by morbidity or mortality). For example, a young patient with a known history of recurrent renal calculi who presents with severe flank pain may be appropriately triaged as high urgency to receive analgesia, but will most likely have a good clinical outcome, whereas an elderly patient with undifferentiated abdominal pain may be triaged as a lower urgency but have higher risk of morbidity and mortality. No triage tool is perfect, and all have issues with sensitivity and specificity resulting in over and under-triage, particularly for certain demographic groups and conditions.^6-8^ There is opportunity to improve triage performance in identifying patients with critical illness, and for improving triage accuracy and the consistency of triage categorisation between healthcare workers.^3^

Machine learning (ML) is a subfield of artificial intelligence (AI), that uses various methods to automatically deduce patterns in data, then makes predictions.^9^ These patterns are learned from the data rather than being explicitly pre-programmed by humans. ML models are iteratively improved through a process called training. In supervised ML training, the model’s predicted output is compared to a “ground truth”, and the error between the predicted value and ground truth is progressively reduced through the training process.^9^ ML models have the potential to improve risk stratification and outcome prediction in the ED setting.^10-12^

Triage has been identified as a promising area to apply ML in the ED.^13, 14^ ML has previously been applied successfully to structured data acquired at triage (such as patient age and vital signs) to predict outcomes including need for admission and intensive care.^15, 16^ Triage nurses routinely collect structured data and an unstructured free-text history of presenting complaint, capturing their impression and subjective assessment about the presentation. This free-text may be more expressive, nuanced, and contain a higher level of information than structured data.^17^ Prior work has suggested that incorporating free-text may improve the performance of ML at ED triage and is an important area for future research despite the challenges of incorporating free-text data into models.^18-20^

Natural language processing (NLP) uses computational methods to analyse and understand human language and its structure.^21^ Early NLP techniques were relatively simple. For example, a “bag-of-words” model bases its decision on the relative frequencies of words in the text, ignoring their order.^22^ These early models often lacked the ability to assess context, negations, and as a result had numerous limitations.^23^ Significant advancements in NLP have been made over the last few years through the use of Deep Learning (DL), a subfield of ML.^24, 25^ DL models pass data through multiple processing layers and in doing so, achieve increasingly abstract representations of the input data, enabling them to learn complex functions.^26^ Massive DL based NLP models have recently been developed.^27-29^ These models have been trained on datasets containing billions of words and have achieved high levels of performance.^27-29^ Some large, pre-trained models, such as Bidirectional Encoder Representations from Transformers (BERT) are publicly available.^27^ Using a pre-trained model allows researchers to take a high performing model as their starting point, and then customise it to their unique needs through fine tuning the model on their local data. For example, Tahayori et al. were able to accurately predict admission from ED using only free-text triage notes and a BERT based NLP model.^30^ Multimodal models integrate NLP with other types of ML to analyse combinations of both free-text data and structured data (such as age and vital signs).

### Objectives

This systematic review aims to evaluate the applications of NLP at ED triage by answering the following questions:

1. How has NLP been applied to ED triage?
2. Do NLP based models outperform humans or current risk stratification techniques?
3. Does incorporating free-text improve predictive performance of ML models when compared to ML models that use only structured data?

## METHODS

A systematic review protocol was prepared in accordance with PRISMA-P guidelines and registered with the International Prospective Register of Systematic Reviews (PROSPERO) on 04/10/2021 (Registration ID: CRD42021276980).^31,32^ All English language peer-reviewed research that applied an NLP technique to free-text obtained at ED triage were eligible for inclusion. As this study aims to broadly assess the capability of NLP at triage, all outcomes and comparators were included. We excluded studies focusing solely on disease surveillance, and studies that used information obtained after triage (such as emergency physician clinical notes and investigations performed within the ED).

We searched PubMed (MEDLINE), Embase, Cochrane Database of Systematic Reviews, Web of Science, and Scopus for research published from 01/01/2012 to present. Electronic databases were first searched on 16/09/2021 and last searched on 01/01/2022. We searched for medical subject headings (MeSH) and text keywords related to NLP and triage. The search strategy was iteratively developed by the multidisciplinary project team that included emergency physicians and computer scientists. The MEDLINE search strategy is provided in Appendix 1, and was adapted to the other databases. Reference lists of the included studies and the authors’ personal archives were reviewed for further relevant literature.

Citations and abstracts were screened independently by two reviewers (JS and JL) against the inclusion and exclusion criteria. Both reviewers were blind to the journal titles, study authors, and institutions. Full text articles were obtained for any articles identified by one reviewer to meet inclusion criteria. Two reviewers (JS and JL) then evaluated the full text reports against the inclusion and exclusion criteria. Data were extracted by JS and JL using a standardised form that included study country, study design, primary outcome, number of sites, study population, input data, NLP and ML models used, comparison, and results. The form was piloted, and calibration exercises were conducted prior to formal data extraction to ensure consistency between reviewers. In cases of conflict or discrepancy, additional review authors were involved until a decision was reached. There were no uncertainties that required authors of the included studies to be contacted.

Data extracted included the study country, study type, outcomes, population, input data, NLP technique, ML method, comparisons, results, public availability of datasets, and public availability of model code. Risk of bias in studies was assessed independently by two authors (JS and JL) using the Prediction model Risk of Bias Assessment Tool (PROBAST).^33^ Due to the high level of heterogeneity between studies, a metanalysis was not conducted. Instead, a narrative synthesis is provided to summarise review findings.

## RESULTS

### Study selection

This process is summarised in a PRISMA Flow Diagram (Figure 1). There were 5099 records identified following database searching and a further 11 records identified through other sources. Following removal of duplicates, 3584 records remained and underwent title and abstract screening. 3448 records were excluded. The remaining 136 full-text articles were assessed for eligibility. In total, 117 articles were excluded, and 19 studies remained for inclusion (Figure 1). There were no unresolved disagreements as to study inclusion or results of data extraction.

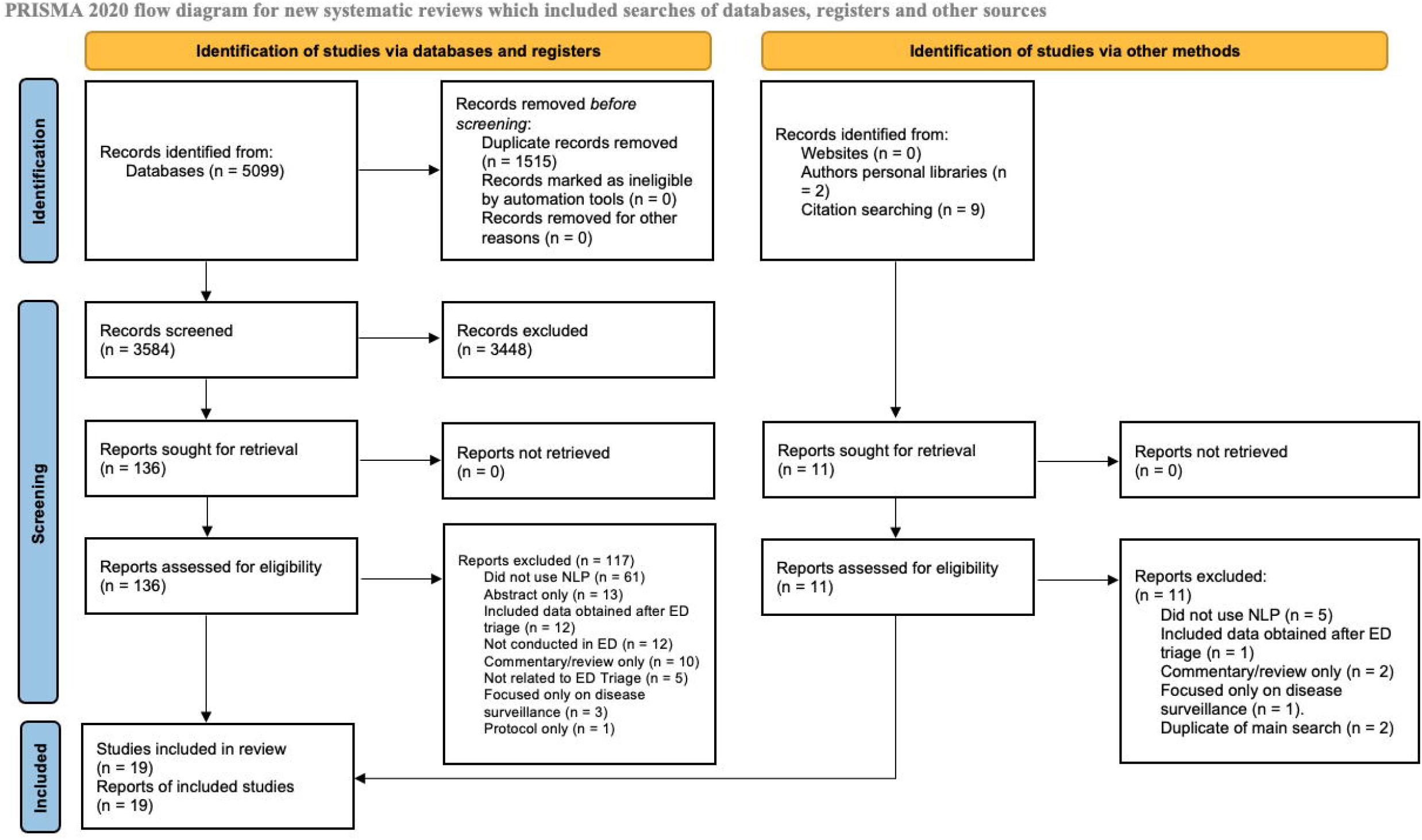

### Characteristics of included studies

A summary of the included studies is shown in Table 1. There were 18 retrospective studies.^17, 18, 30, 34-48^ One study reported their ML model was developed using retrospective data then validated using prospective data.^49^ All used observational cohort designs. Two studies were international multi-centre studies (USA and Portugal); 11 were conducted in the USA; 2 were from South Korea; one each from Australia, Brazil, China, and France. The most common primary outcomes assessed were prediction of triage score (six studies), prediction of admission (five studies), and prediction of critical illness (three studies). Two studies predicted need for imaging within the ED, two studies looked at the assignment of provider assigned chief complaint label, and one study predicted diagnosis of infection in the ED.

**Table 1.**
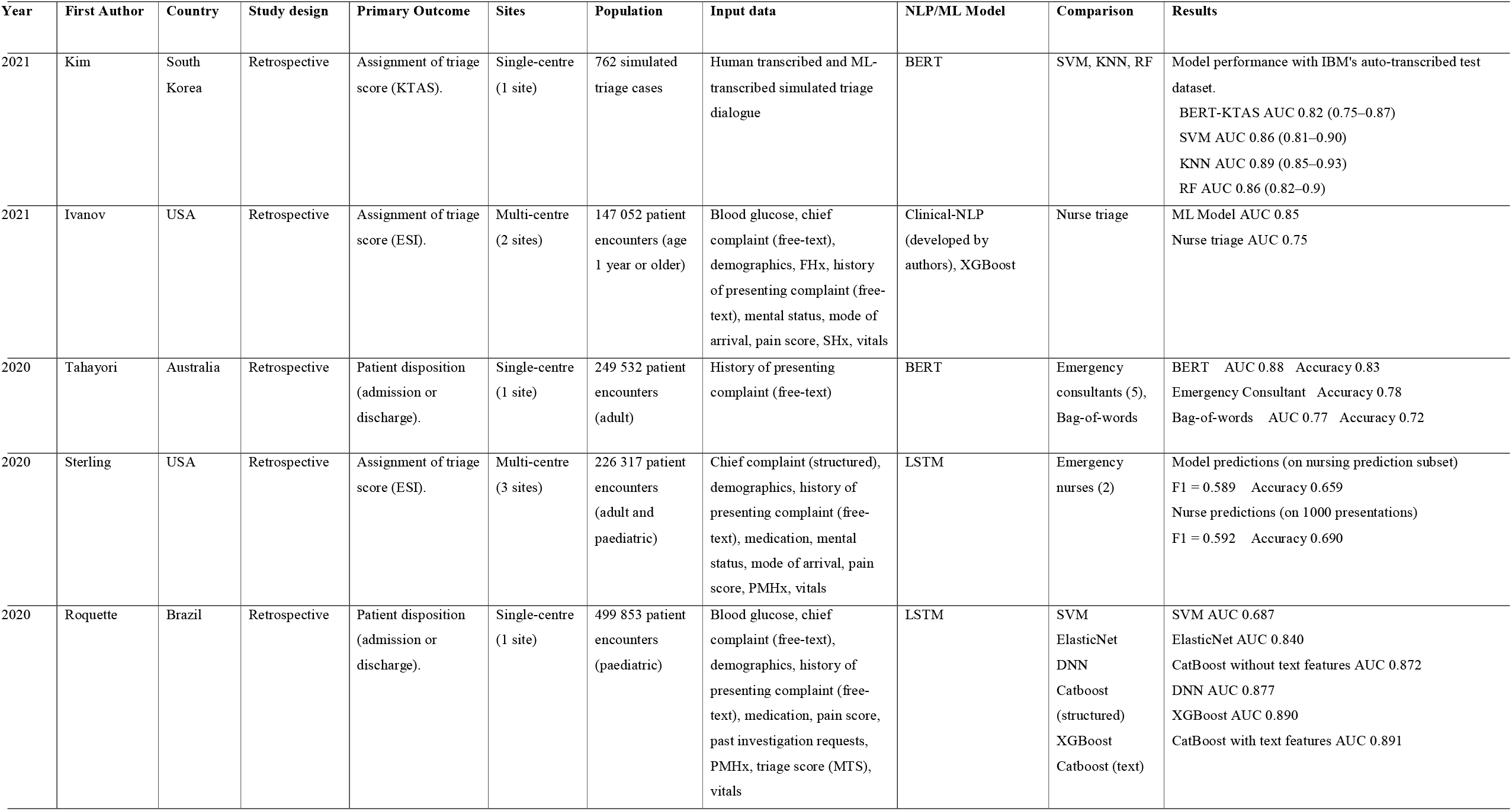
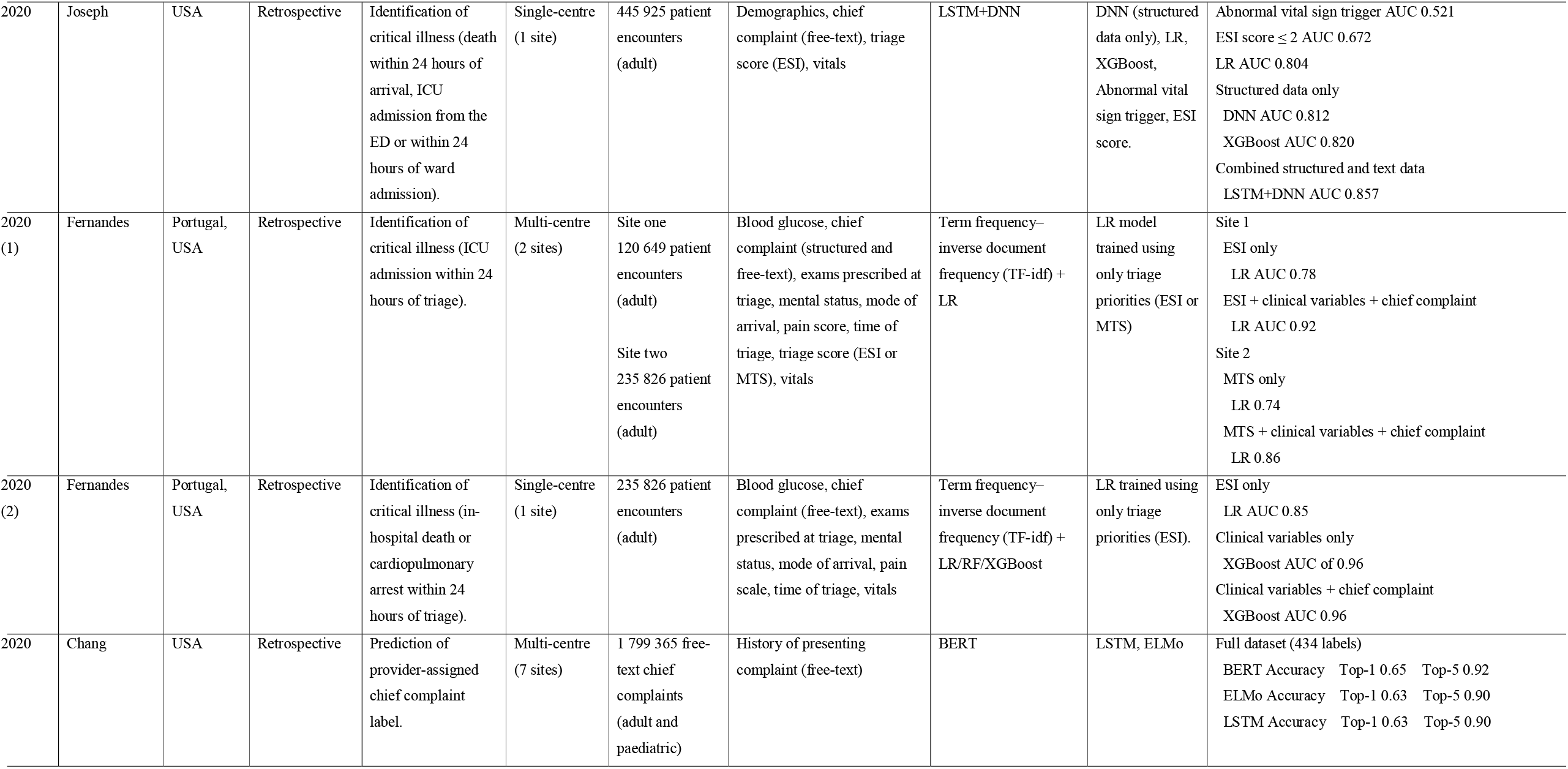
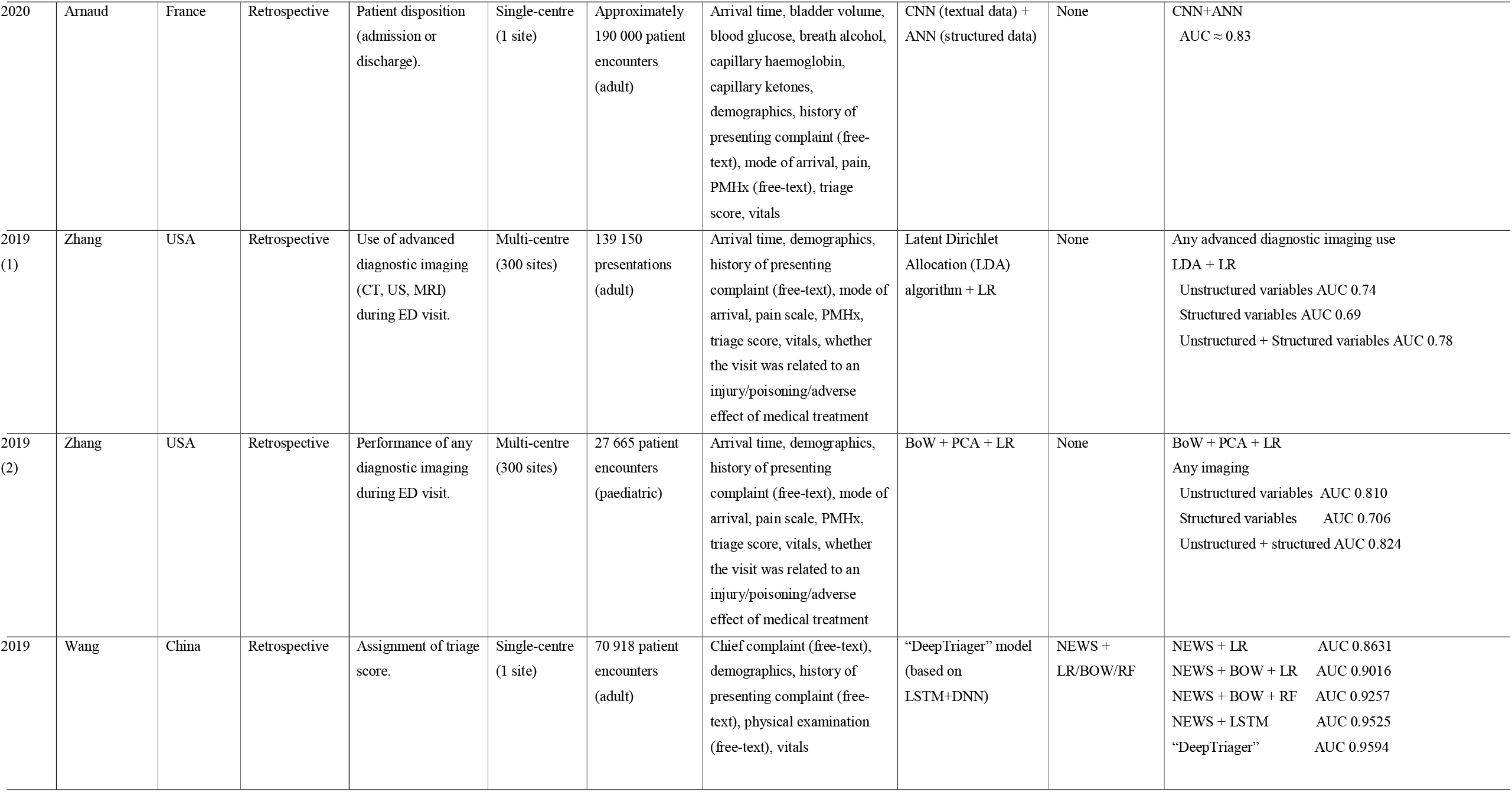
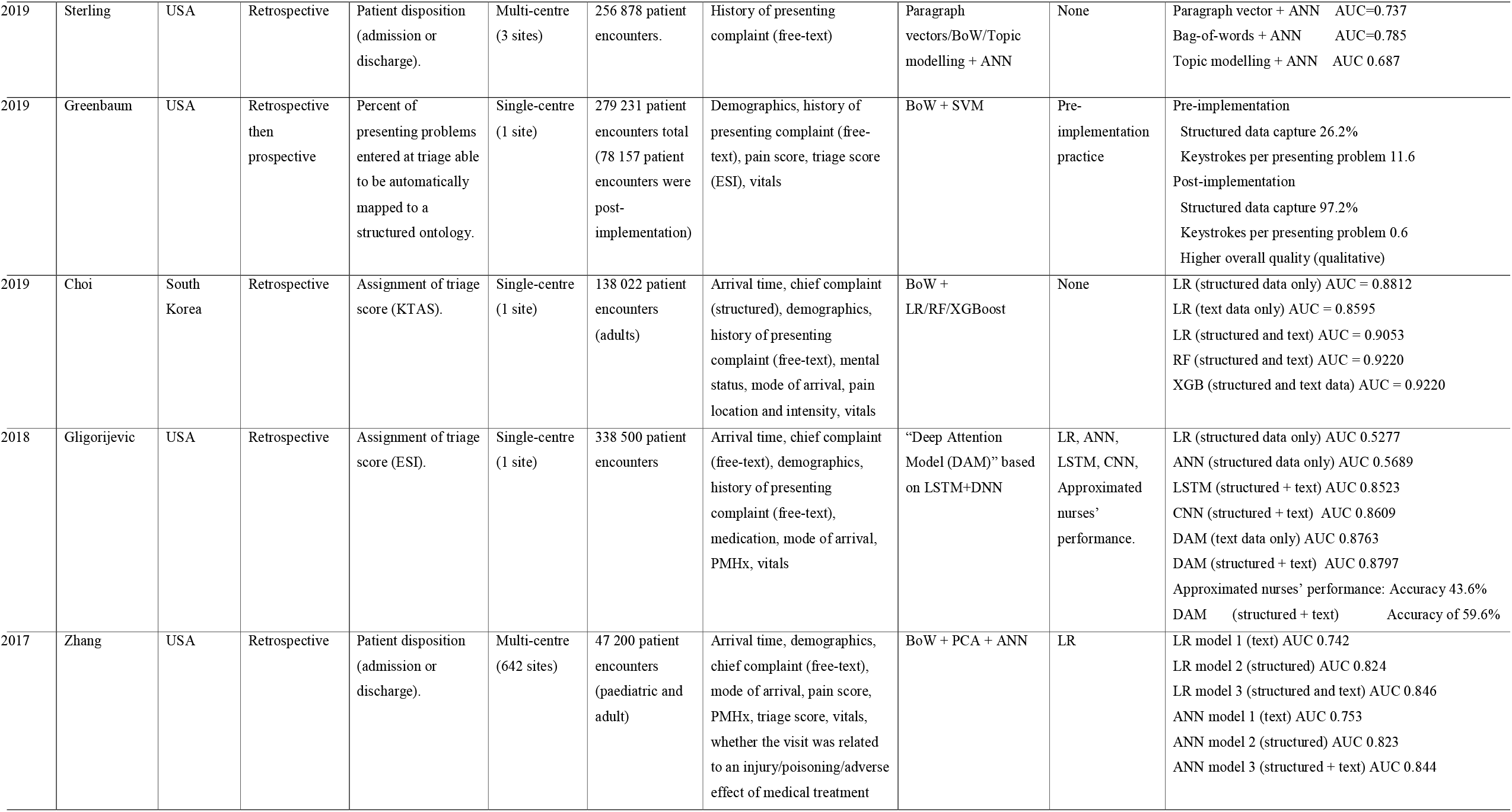
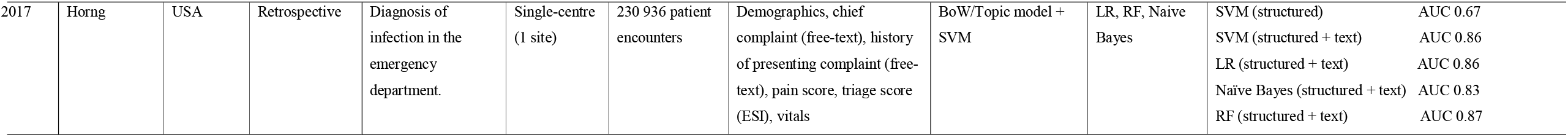
Summary of included studies. Abbreviations KTAS - Korean Triage and Acuity Scale ESI - Emergency Severity Index ICU - Intensive Care Unit ED - Emergency Department ML - Machine learning FHx - Family history SHx - Social history PMHx - Past medical history Vitals - Respiratory rate (RR), heart rate (HR), systolic blood pressure (SBP), diastolic blood pressure (DBP), temperature (Temp), and oxygen saturation (SPO2). MTS - Manchester Triage system BERT - bidirectional encoder representations from transformers XGBoost - eXtreme Gradient Boosting LSTM - Long short-term memory DNN - Deep neural network LR - Logistic regression RF - Random forest CNN - Convolutional neural network ANN - Artificial neural network BoW - Bag-of-words PCA - Principal component analysis SVM - Support vector machine KNN - k-nearest neighbors F1 - the harmonic mean of precision and recall AUC - Area under the receiver operating characteristic curve ELMo - Embeddings from Language Model NEWS - National Early Warning Score

The population size varied greatly between studies ranging from 1.8 million patients to 762 simulated encounters. Four studies used a population of under 100 000, four studies had a population of between 100 000 and 200 000, six studies had a population of between 200 001 and 300 000, and six studies had a population of over 300 000. Eleven studies used data from a single site and eight studies used data from multiple sites. The largest number of sites used was 642 by Zhang et al.

Fourteen studies applied NLP to free-text history of presenting complaint, seven studies applied NLP to a free-text chief complaint, two studies applied NLP to a structured chief complaint label, and one study applied NLP to simulated triage dialogues that had been transcribed by either a human or an ML model. The other most frequently used input variables were patient demographics (13 studies), patient vital signs (heart rate, respiratory rate, oxygen saturation, blood pressure, and temperature) (15 studies), pain score (12 studies), triage score (10 studies), mode of arrival (10 studies), time of arrival (8 studies) and past medical history (7 studies). Other input variables included mental status (5 studies), and blood glucose level (5 studies).

### Prediction of admission

NLP models and multimodal models were able to accurately predict admission at time of triage for adult and paediatric patients.^18, 30, 35, 41, 46^ Of the five studies focusing on predicting admission to hospital, Roquette et al. achieved the highest Area Under the Receiver Operating Characteristic Curve (AUC) using a gradient boosting model (AUC 0.89). Tahayori et al. achieved a similar AUC (0.88) using only free-text history of presenting complaint. Tahayori et al. were the only authors that compared their model to emergency physician performance. Their model achieved a higher accuracy than five emergency consultants (0.83 vs 0.78) and higher specificity (0.86 vs 0.77), but lower sensitivity (0.72 vs 0.9). Roquette et al. and Zhang et al. both compared ML models trained using structured data only with ML models that incorporated both structured data and text data. They found that the addition of text data results in a small improvement when compared to the use of structured data alone.

### Prediction of critical illness

Multimodal models were able to accurately predict critical illness in adult patients, defined as ICU admission, cardiopulmonary arrest within 24 hours, or death within 24 hours of triage.^43-45^ Of the three studies that predicted critical illness at triage, Fernandes et al. achieved the highest AUC (0.96) in predicting in-hospital death or cardiopulmonary arrest within 24 hours of triage using an extreme gradient boosting model. They found no difference in AUC when using clinical variables only or clinical variables and structured chief complaint processed by NLP. Joseph et al. found their NLP model (AUC 0.857) significantly outperformed an abnormal vital sign trigger (AUC 0.521) and ESI score ≤ 2 (AUC 0.672) in predicting critical illness. The addition of free-text data improved the performance of their neural network model (from AUC 0.820 to AUC 0.857).

### Prediction of triage score

NLP has been applied in multiple triage systems. NLP models and multimodal models were able to accurately assign triage categories using structured and free-text data.^17, 36^-^38, 47, 48^ Wang et al. achieved the highest performance in predicting ESI using their “DeepTriager” model (AUC 0.96). Kim et al. achieved an AUC of 0.89 in assigning a KTAS category to auto-transcribed simulated triage dialogue. This was only slightly lower than the performance achieved using human-transcribed simulated triage dialogue (AUC 0.90). Three studies compared the accuracy of triage scores assigned by multimodal models incorporating NLP to triage scores assigned by nurses.^17, 36, 47^ Such models were more accurate than nurses in two out of three papers.^36, 47^ The addition of text data compared to structured data alone improved performance in assigning triage score.^36, 37^

### Prediction of provider-assigned chief complaint

NLP models and multimodal models incorporating NLP were able to accurately map free-text history of presenting complaint to structured chief complaints.^42, 49^ Chang et al. (2020) used BERT to accurately predict provider-assigned chief complaint labels (Top-5 structured label AUC 0.92). Greenbaum et al. (2019) iteratively developed their own structured ontology and were eventually able to map 97.2% of presentations to their structured ontology using their NLP based predictive model.

### Prediction of investigations

Multimodal models incorporating NLP were able to predict diagnostic imaging performed in the ED.^39, 40^ Zhang et al. developed a model to predict need for advanced diagnostic imaging (computed tomography, ultrasound, magnetic resonance imaging) in the ED, and obtained an AUC 0.78 using a “bag-of-words” model. Zhang et al. were also able to predict the need for any diagnostic imaging in a paediatric population with an AUC 0.824. The inclusion of unstructured variables improved performance slightly in both cases.

### Identifying infection

Horng et al. (2017) found that the incorporation of free-text data improves the discriminatory ability (increase in AUC from 0.67 to 0.86) for identifying sepsis (defined by ICD-9-CM code) in the ED at triage.

### Multimodal models

Eleven papers compared ML models that used only structured data to multimodal models that incorporated both structured data and free-text data.^34-40, 43-46^ The best performing model in each of these papers incorporated free-text. The largest improvement in model performance from incorporating free-text was found by Horng et al. (increase in AUC from 0.67 to 0.86 for identifying infection). The addition of free-text did not improve model AUC in one case, however, did improve model average precision.^44^ There were no cases where the incorporation of free-text into the model resulted in worse performance. Six papers assessed models that used only free-text, with no structured data.^30, 36, 37, 39, 40, 42^ Tahayori et al. were able to use only free-text data to predict admission with high accuracy (83%). Zhang et al. used free-text to predict performance of diagnostic imaging. Gligorijevic’s “Deep Attention” models using only unstructured data outperformed those using only structured data. Incorporating both structured data and free-text data improved results when compared to models that used only free-text data, though often only a small improvement was found.

### Modern NLP compared to traditional NLP

Three papers directly compared modern NLP based on DL to more traditional ML techniques such as bag-of-words and topic modelling.^30, 38, 48^ Modern DL based NLP outperformed traditional ML based NLP in two cases.^30, 38^ In contrast, Kim et al. found that a BERT based DL model did not perform better than ML based models, though their population was relatively small. Chang et al. compared the performance of multiple modern DL based models, finding BERT slightly outperformed Embeddings from Language Models (ELMo) and Long Short-Term Memory (LSTM) networks in mapping free-text chief complaints to structured fields.

### Integration into practice

Greenbaum et al. was the only study that reported the deployment of an NLP based model into clinical practice. Greenbaum et al. aimed to increase the ease of high-quality structured data collection at triage through the use of an NLP based model. Their model used both free-text triage notes and structured data to provide contextual autocomplete of chief complaint label, and also show the user a list of the top five most likely chief complaints. Prior to implementation of their model, 26.2% of patient encounters resulted in structured data capture. Following implementation this increased to 97.2%. The authors aggregated multiple incidents of unscheduled downtime that occurred throughout the study to assess the impact of their model. When ML based autocomplete was not operational (and instead alphabetised autocomplete was shown), the percent of encounters that resulted in structure data capture decreased from 97.2% to 89.2%. The number of keystrokes typed for each presenting problem decreased from 11.6 pre-implementation to 0.6 post implementation. Contextual autocomplete was associated with qualitatively more complete and higher quality structured documentation of chief complaints.

### Study quality—Risk of bias within and across studies

A summary of the PROBAST assessment is provided in Table 2. Overall, 15 studies were considered to have a high risk of bias. Four studies were assessed as having a low risk of bias. One study had high applicability concerns and 18 studies had low applicability concerns. The four studies assessed as having low risk of bias also had low applicability concerns. No studies referred to a previously published or publicly registered protocol.

**Table 2.** PROBAST assessment of the included studies. PROBAST = Prediction model Risk Of Bias ASsessment Tool; ROB = risk of bias. + indicates low ROB/low concern regarding applicability; − indicates high ROB/high concern regarding applicability; and ? indicates unclear ROB/unclear concern regarding applicability

| Study | Risk of Bias (ROB) |  |  |  | Applicability |  |  | Overall |  |
| --- | --- | --- | --- | --- | --- | --- | --- | --- | --- |
|  | Participants | Predictors | Outcome | Analysis | Participants | Predictors | Outcome | ROB | Applicability |
| Kim et al. 2021 | ? | ? | + | - | - | - | + | - | - |
| Ivanov et al. 2021 | + | + | - | - | + | + | + | - | + |
| Tahayori et al. 2020 | + | + | + | + | + | + | + | + | + |
| Sterling et al. 2020 | ? | + | ? | - | + | + | ? | - | + |
| Roquette et al. 2020 | + | + | + | + | + | + | + | + | + |
| Joseph et al. 2020 | + | + | + | - | + | + | + | - | + |
| Fernandes et al. 2020 (2) | - | + | ? | + | + | + | + | - | + |
| Fernandes et al. 2020 (1) | - | + | ? | + | + | + | + | - | + |
| Chang et al. 2020 | + | + | - | + | + | + | + | - | + |
| Arnaud et al. 2020 | - | + | + | - | + | + | + | - | + |
| Zhang et al. 2019 (2) | ? | + | + | - | + | + | + | - | + |
| Zhang et al. 2019 (1) | ? | + | + | - | + | + | + | - | + |
| Wang et al. 2019 | + | + | + | - | + | + | + | - | + |
| Sterling et al. 2019 | + | + | + | - | + | + | + | - | + |
| Greenbaum et al. 2019 | + | + | + | + | + | + | + | + | + |
| Choi et al. 2018 | + | + | ? | - | + | + | + | - | + |
| Gligorijevic 2018 | + | + | - | ? | + | + | + | - | + |
| Zhang et al. 2017 | + | + | + | + | + | + | + | + | + |
| Hornig et al. 2017 | + | + | - | + | + | + | + | - | + |

### Availability of datasets and code

Availability of study datasets and code is shown in Table 3. Data was publicly available for three studies (all by Zhang et al.) and was available on request from study authors for a further four studies.^30, 34, 35 39, 40 43, 44^ One study reported plans to release a modified de-identified dataset, however at the time of this review this is still pending approvals.^45^ The model code was publicly available for two studies.^42, 45^ Notably, the code repository from Chang et al. was well organised and contained clear instructions for researchers on how to download their pretrained model and apply it to their own dataset.

**Table 3.** Availability of dataset and code for included studies. * Data available on request from the authors and may be released to researchers following the signing of a data sharing agreement. ** Pending approval, a modified, de-identified dataset containing modified chief complaint text data will be uploaded. Approval still pending at time of this review. *** All data freely and publicly available.

| **Study** | **Dataset available** | **Code available** |
| --- | --- | --- |
| Kim et al. 2021 | No | No |
| Ivanov et al. 2021 | No | No |
| Tahayori et al. 2020 | Yes* | No |
| Sterling et al. 2020 | No | No |
| Roquette et al. 2020 | No | No |
| Joseph et al. 2020 | No** | Yes |
| Fernandes et al. 2020 (2) | Yes* | No |
| Fernandes et al. 2020 (1) | Yes* | No |
| Chang et al. 2020 | No | Yes |
| Arnaud et al. 2020 | No | No |
| Zhang et al. 2019 (2) | Yes *** | No |
| Zhang et al. 2019 (1) | Yes *** | No |
| Wang et al. 2019 | No | No |
| Sterling et al. 2019 | No | No |
| Greenbaum et al. 2019 | No | No |
| Choi et al. 2018 | No | No |
| Gligorijevic 2018 | No | No |
| Zhang et al. 2017 | Yes*** | No |
| Hornig et al. 2017 | Yes* | No |

## DISCUSSION

### NLP at triage

This review finds that NLP has been applied to data available at the time of ED triage to predict a range of outcomes, with a focus on predicting need for admission and assigned triage score. The results of this review also highlight that unstructured free-text triage notes contain valuable information. Through NLP techniques, this information has started to become accessible to use for automated predictive purposes. The combination of free-text nursing triage notes with structured data appears to result in the best model performance, however free-text nursing triage notes alone can be used by NLP algorithms to predict need for admission and need for diagnostic imaging.^18, 30, 39, 40^ A benefit of developing models that require only free-text as an input is that it may allow for easier portability of predictive models between different triage systems.^30^

### Structured data capture

Accurate and consistent structured capture of patients’ presenting complaints is important for research, service improvement, and public health initiatives.^49^ Common medical ontologies also improve system interoperability.^50^ However collection of structured data is often difficult, especially when contrasted with the ease and expressiveness of free-text entry.^49^ In a rare singular example of NLP being deployed into routine clinical practice at ED triage, Greenbaum et al. developed, implemented, and prospectively evaluated an NLP driven user interface to mitigate the challenges of structured data capture.^49^ Promisingly, they report that their NLP based contextual auto-predict did not add additional burden to users and made structured data collection easier than unstructured data collection. Because of this, structured data collection increased significantly.

### Improving ED workflow and efficiency

ED overcrowding is a serious issue worldwide, with significant negative impact on patient morbidity and mortality. Having an emergency physician triage patients (or implementing a rapid assessment zone) enables early senior clinician input and decision making, and can lead to a reduced patient ED length of stay.^51, 52^ Patient time spent in the waiting room is likely underutilised.^52^ NLP could be applied to triage notes to predict which patients will likely require investigations such as blood tests or imaging, and in doing so allow for these investigations to be ordered immediately on arrival, rather than only being ordered after they are seen by a doctor. An emergency physician could review and then approve or reject suggested investigations. In this way, applying NLP to triage could leverage the expertise of the emergency physician.

Delays in specialist consultation and subsequent specialist review contribute to reduced ED throughput, and improvements in the consultation process from the ED have the potential to reduce ED length of stay.^53^ Using NLP to identify at the point of triage, patients who are likely to require admission could assist with hospital resource allocation, improve patient flow, and allow for anticipation of system stressors, such as worsening access block.^18, 30^ Bed allocation could begin at the time of patient triage, rather than hours into a patient’s ED stay.^30^ To fully realise the potential of predicting admission at triage, the NLP model would need to be supported by other infrastructure. For example, an “early admission team” could review patients who are flagged as very likely to be admitted, or stable patients not needing acute resuscitation could be diverted away from the ED and sent to the appropriate specialty team.

### NLP compared to humans

Human performance may be a reasonable baseline for ML models to meet to be considered accurate enough for implementation into clinical practice. Few studies have compared NLP models at triage to human performance. Such comparisons will be crucial in future work.

Tahayori et al. was the only study that compared results from NLP models to emergency physicians.^30^ Ivanov et al., Sterling et al, and Gligorijevic et al. compared NLP based models to nurses in assigning triage scores and found model accuracy was similar to nurses.^17, 36, 47^

### Interpretability

Few papers attempted to address human interpretability of models. While DL has been criticised as being a “black box”, there is ongoing work to develop more “explainable AI”.^54, 55^ Wang et al. show how models could be somewhat more interpretable.^38^ Their triage model is able to highlight free-text triage notes, with a darker colour corresponding to the sections of text that was more heavily weighted by the model. This provides an initial “sense check” that humans can then combine with their own experience and knowledge.

### Modern NLP

While it is difficult to compare studies due to their heterogeneity, advanced DL based NLP appears to outperform traditional NLP. This is certainly the case when compared internally within studies and is consistent with previous NLP research.^56^ BERT appears to be the most popular advanced NLP that has been used. BERT was released in October 2018 and at the time of release, BERT outperformed other NLP models.^27^ However, of the 16 papers published since the release of BERT, only three have used it. Other large models have subsequently been released. For example, GPT-3 is a 175 billion parameter language model that was released in 2020 and is reported to outperform BERT in various circumstances.^28^ Chowdhery et al. have recently published Pathways Language Model (PaLM), a 540-billion parameter model that achieves further increases in performance.^29^

## FUTURE DIRECTIONS

Triage is a promising place to start applying NLP in the ED. Large datasets with clearly labelled outcomes makes triage well suited to applications of ML. Triage information is often available hours before emergency physician documentation, and accurate predictions made at triage have the potential to increase healthcare system efficiency.^18^ There is also the possibility of close human oversite if deployed in practice. Future work could aim to predict other important patient-oriented outcomes at the time of triage such as wait times, need for advanced cardiovascular investigations, or need for surgery.

### Incorporating clinical gestalt

Sterling et al. 2020 noted the difficulty in capturing the general clinical impression of the triage nurse.^17^ Ivanov et al. also noted that important contextual aspects at triage were not available for consideration by ML models.^47^ Future work could assess the impact of incorporating triage nurses’ gestalt into predictive models. This could be expanded to also capture patients’ predictions regarding their need for admission to hospital. Other contextual data available at the time of triage such as the number of patients currently waiting, the number of patients currently in the ED, and number of admitted patients in the hospital could also be incorporated into ML models.

### Integration with other AI systems

There are opportunities to integrate NLP as part of a larger AI based system. Kim et al. provides an interesting example of how various AI based technologies can be combined.^48^ Speech recognition could be used to automatically generate a transcript of the entire triage conversation, which could then be used by NLP models. However, the performance of speech recognition technologies would likely deteriorate in a noisy ED, and combining multiple complex AI based technologies raises the possibility that small initial errors could be amplified as they propagate through the models. NLP models at triage could also be integrated with other novel AI based interventions, such as automated monitoring of patients’ vital signs while they are in the waiting room, or with data entered by patients themselves in AI based self-triage applications.

### Pre-trained models for ED triage

Publicly available large DL based language models have often been trained on corpuses containing text from newspapers, books, and websites.^27, 28^ Triage notes are often quite short and contain a number of unique and idiosyncratic abbreviations and acronyms not common in everyday English language.^17, 30^ The benefits of applying DL based NLP models to triage notes may yet to be fully realised, as they were not developed for triage specific purposes. DL based NLP models that have been fine-tuned on large corpuses of medical text have been released, however they have not been applied to ED triage. Large publicly available clinical databases such as MIMIC-IV that contain ED triage notes with linked outcomes are likely to be helpful in further model development and may facilitate direct comparisons between models developed by different research groups.^57, 58^ Triage focused NLP research could also benefit from groups sharing large language models that have been pre-trained on triage data. These models could be used as starting points by others, though it is unknown whether such models’ performance would generalise across different healthcare settings and triage systems. It is also unknown if the length of triage notes impacts model performance. This could be evaluated in future work.

### Prospective and external validation is needed

The majority of research so far has been retrospective and completed in the USA. There is a significant need for prospective evaluation and external validation, especially in other countries and triage systems.

### Clinical impact and risk

NLP models have rarely been deployed at ED triage. As such, it is unknown what impact these tools could have on clinical practice. The introduction of a new tool into a complex system is likely to have unintended consequences, and use of the tool may itself change practice. Triage notes may be written in a different way if it is known that they are being used for predictive purposes. There may also be unintended harms. For example, telling a patient at triage that they are likely to be admitted or to have a long wait time, could influence their behaviour and increase the number of patients who leave without being seen. It may be useful to establish the performance benchmarks predictive models must meet prior to implementation into clinical practice. This could be done through further studies comparing NLP model performance to emergency physicians and nurses. Further research is also required to understand how to best integrate early admission predictions into hospital systems and clinical practice.

NLP models can be retrained and updated as new data becomes available. Therefore, model performance may change over time. It will be important to ensure that there is appropriate algorithm stewardship in place prior to clinical use.^59^ Predictive models are trained on data that reflects current practice. This engrains the assumption that current practice is appropriate, which may not be the case.

### Acceptability

It is also unknown if the use of NLP at triage is acceptable to patients and staff. It will be important to involve clinicians, patients, and healthcare consumer groups in the development and governance of any future implementation projects. It will also be important to ensure that these systems do not place further burden on users. Ease of use and perceived clinical impact will likely be important factors for adoption by clinicians.

### Ethical issues

Race, age, and gender biases at ED triage have been previously reported.^60-62^ Concerns over bias in ML models have been well described, and new tools are being developed to assess such biases.^63-65^ At its best, NLP at triage could help reduce bias through standardising triage decisions and providing a more objective triage score. However, at its worst NLP at triage could further ingrain existing biases into practice, under the guise of objectivity and hidden in the opacity of abstract algorithms. Patient apprehensions and concerns about the use of AI will also need to be considered. While the emerging body of literature shows patients view AI largely positively, they do have some concerns with its use in healthcare.^66^ These include perceptions that AI is less accurate than clinicians, there is a lack of transparency in predictions, and there are risks to the privacy of their personal healthcare data.^67-72^ Further research investigating the impact of NLP based tools on vulnerable and minority populations is warranted.

## LIMITATIONS

### Study level

Only one study contained prospectively validated results, and no studies contained results that were externally validated at a separate site. Results reported may not be generalisable to other settings. There was inconsistent reporting of methods and results among studies. The majority of studies (79%) were assessed to have a high risk of bias.

### Review level

Heterogeneity of the included studies precluded meta-analysis which limits the level of evidence this review provides. All studies reported positive results for NLP at triage, which may reflect publication bias. While we took significant care to ensure our search strategy was broad enough to capture all relevant literature, the variety of NLP and ML terminology means that some studies may have been missed. Non-English articles, and articles published prior to 2012 were also excluded from our search.

## CONCLUSION

The use of NLP at triage appears feasible and could accurately predict important patient-oriented outcomes including need for admission and need for critical care. However, there are few examples of implementation into clinical practice and most research is retrospective. The potential benefits of using NLP at triage are yet to be realised. Further research is needed to prospectively assess the acceptability and impact of implementing NLP at triage on staff, patients, and the healthcare system.

## Supporting information

PRISMA Abstract Checklist

PRISMA Checklist

## Data Availability

All relevant data are within the manuscript and its Supporting Information files.

## Competing Interested

No authors declare any competing interests.

