## Appendix 1 for "Applications of Natural Language Processing at Emergency Department Triage: A Systematic Review"

**S1 Appendix – Medline Search Strategy**

| 1 | "natural language processing".tw. |
| --- | --- |
| 2 | "nlp".tw. |
| 3 | exp Data Mining/ |
| 4 | "data mining" .tw. |
| 5 | "text mining".tw. |
| 6 | exp Artificial Intelligence/ |
| 7 | "artificial intelligence".tw. |
| 8 | "machine learning*.tw. |
| 9 | "deep learning".tw. |
| 10 | exp Triage/ |
| 11 | "triage".tw. |
| 12 | 1 or 2 or 3 or 4 or 5 or 6 or 7 or 8 or 9 |
| 13 | 10 or 11 |
| 14 | 12 and 13 |
| 15 | limit 14 to yr="2011 -Current" |
